# Developing a Community Engaged Sickle Cell Disease Center: An Initial Data Analysis of Healthcare Utilization Rates and Perspectives of Key Informants

**DOI:** 10.1101/2021.04.07.21254482

**Authors:** Hannatu Tunga-Lergo

## Abstract

**Background:** The mortality rate of individuals with Sickle cell disease (SCD), the most prevalent genetic disease in the United States, has been increasing at 1% per year. It has been declared a global and national public health priority by the World Health Organization (WHO) and Centers for Disease Control and Prevention (CDC). As a complex chronic and acute condition, preventive care and patient management of SCD requires a patient-centered, comprehensive, and multidisciplinary approach; unfortunately, few SCD treatment centers use this approach. Moreover, individuals with SCD are at the intersectionality of race and socioeconomics and thus face additional barriers to access to quality care, which may ultimately result in higher utilization of acute care services, especially during the transitioning period from pediatric to adult care. Greater acute care utilization has been found to be associated with higher mortality rate and severely compromised health related quality of life; thus, it is important to assessing needs of SCD patients as they relate to access to quality care.

**Objective:** The aim of this study was to conduct a preliminary needs assessment for the development of a community engaged SCD center. This study also aimed to determine if frequency of acute care utilization was associated with age and insurance type, to provide surveillance data, and to identify opportunities to address barriers to access to quality care from key informant (local and cross-institutional) perspectives.

**Method:** A retrospective cohort study of SCD related emergency department (ED), inpatient hospitalization, and outpatient clinic utilization encounters, which occurred from 09/01/2012-06/01/2019, was queried from UF Health’s Integrated Data Repository (IDR). Quantitative analysis, frequencies, proportions, and Pearson Chi-square inference were conducted on the administrative data received. Further, key informant interviews of stakeholders in Alachua County, FL and Yale New Haven Health’s Adult Sickle Cell Program, New Haven, CT were performed. An iterative qualitative thematic analysis of their perspectives was conducted.

**Result:** There were 27,932 total encounters that were stratified by age and payer type. The average length of hospitalization stay was .71 ± 3.84. The 18-30-year-olds had the highest proportion of ED utilization (34.7%), hospitalizations (32.1%), and outpatient clinic utilization (26.4%). This was followed by the 31-45-year-olds with 20.4% of ED utilizations, 22% of hospitalizations, and 20.5% of outpatient clinic utilizations. Those with public health insurance accounted for 74% of ED encounters, 81% of hospitalizations, and 82% of outpatient encounters. Common themes and subthemes from key informant interviews included: champion, transition of care, pain management, bias, patient and family education, provider knowledge, social worker, multidisciplinary/comprehensive care, mental health, education, and employment.

**Conclusion:** Among adults with SCD in the UF Health system, younger adults (e.g., those who are transitioning into adult care) and those with public insurance utilized acute care services at greater proportions, indicating a need to identify and address possible barriers to access to quality care.

---

Sickle Cell Disease (SCD) is a chronic genetic disorder that is a globally and nationally recognized health disparity with a devastating sequalae. It was first discovered in 1910, by Dr. Herring, from the observation of “sickle” shaped cells in a man suffering from severe pain episodes and anemia (Herrick, 2001). In 1951, the Nobel Prize Laureate Linus Pauling and his colleague Dr. Harvey Itano coined the term “molecular disease” from the discovery of the different chemical structures of hemoglobin in SCD patients (Pauling, Itano, Singer, & Wells, 1949). SCD is the first discovered and longest known genetic disease (Winter, n.d.) which pioneered molecular medicine and was “at the leading edge of investigations to elucidate the molecular basis of human diseases” (Frenette, & Atweh, 2007, p.850).

SCD is an autosomal recessively inherited genetic hemoglobinopathy characterized by a mutation on the sixth residue of the β-globin polypeptide which results in a substitution of valine for glutamine. There are different genetic variants, with the homozygous HbSS being the most common and severe form. HbSC is typically mild and HbSβ+-thalassemia varies depending on the HbSβ+-thalassemia severity. As such, the HbSβ+-thalassemia variant can be mild, as commonly found in people of African descent, or moderate, as commonly found in people of Mediterranean descent. However, the HbSβ0-thalassemia variant tends to be severe, similar to HbSS (Frenette, & Atweh, 2007).

SCD is the most common genetic disease in the world and is a global public health priority that still has unmet needs (McGann, 2016). It affects individuals of all races and ethnicities (Ojodu, Hulihan, Pope & Grant, 2014). It predominantly tends to be globally distributed in tropical areas such as Africa, India, Middle East, and the Mediterranean and with migration can also be found in North America, Central America, South America, Northern Europe, and Australia. Stemming from the evolutionary origins of SCD as a protective mutation to defend against malaria (Sabeti, 2008). Epidemiologically, the exact prevalence of SCD is unknown. It is estimated that globally the number of newborns with SCD will increase from 305,800 in 2010, to 404,200 by 2050 (Piel, Hay, Gupta, Weatherall & Williams, 2013). In the United States, SCD is also the most common genetic disease and it is estimated that approximately 100,000 Americans have SCD (Hassell, 2010) and 2 million have the trait. It is estimated that in the state of Florida it affects 8,374 to 14,236 African Americans (Walker, Brown & Lopez, 2014).

The genetic point mutation, where the amino acid valine replaces glutamic acid causes an atypical β hemoglobin gene. As a result, when in a deoxygenated state this substituted hemoglobin polymerizes and aggregates into a stiff and inflexible sickled shaped red blood cell (RBC), making it difficult to carry oxygen. Additionally, the shape makes it difficult for the RBC to travel through capillaries to deliver oxygen. They are also more fragile resulting in RBCs that live only 10-20 days and frequently hemolyze, leading to chronic anemia (Quinn, 2013). The sickle cells also have a stickiness property to them. They adhere to the vascular endothelium and create microvascular clumping of cells and occlusion of blood vessels. This leads to painful vaso-occlusive ischemic and hypoxic episodes (Hebbel, et. al, 1980), called a crisis. A crisis, predominantly defined by severe pain, is the most debilitating and most common reason for acute care utilization (Emergency department (ED)) visits (Sedrak, & Kondamudi, 2018).

As SCD is a blood disorder, it affects every single part of the human body. Due to chronic damage from crises some lose organ/organs in their entirety. In particular, most lose their spleen, or must have it removed due to splenic sequestration and auto-infarction. As a result, they are also categorized as immunocompromised patients and must remain vigilant to get immunizations and to avoid infections (a leading cause of death) from their social and physical environment (Ammann, Addiego, Wara, Lubin, Smith, & Mentzer, 1977). Symptoms can also include anemia, fatigue, pulmonary hypertension, kidney and liver problems, joint necrosis, eye problems, jaundice, etc. with acute chest syndrome and stroke as additional leading causes of death (Sedrak, & Kondamudi, 2018; NIH What is Sickle Cell Disease).

In addition to physical complications, social determinants have a significant impact on the health and quality of life (QOL) of individuals with SCD. Individuals with SCD may experience negative psychosocial effects of their disease condition (Vilela, Cavalcante, Cavalcante, Araújo, Lôbo& Nunes, 2012), which diminishes their overall QOL, in a proportionally inverse relationship to the severity of the disease (Pereira, Brener, Cardoso & Proietti, 2013). QOL, as defined by the World Health Organization (WHO), is “an individual”s perception of their position in life in the context of the culture and value systems in which they live and in relation to their goals, expectations, standards and concerns” (Whoqol Group, 1995). SCD negatively impacts their physical, psychological, social, and occupational well beings, as well as their “levels of independence and environment” (Thomas & Taylor, 2002).

The QOL of individuals with SCD is also diminished as SCD is a complex, acute and chronic, rare blood disease where pain is the norm. Pain is debilitating, recurrent, and unpredictable (Howard, Thomas, & Rawle, 2009). In addition, access to healthcare is impaired, further diminishing their health and QOL (Dos Santos & Gomes Neto, 2013). Painful crisis episodes involve frequent hospitalizations and many sick days (Farber, Koshy, Kinney, & Cooperative Study of Sickle Cell Disease, 1985) and leads to “social and financial crises” (Bhagat, Baviskar, Mudey, & Goyal, 2014). Missed days in school affects the chronicity of educational attainment, while missed work affects their long-term employment prospect. Thus, the majority of individuals with SCD become unemployed, have reduced productivity, become reliant on government subsistence, and move down the socioeconomic ladder (Farber, Koshy, Kinney, & Cooperative Study of Sickle Cell Disease, 1985). SCD additionally impairs their social networks as they are more likely to report higher levels of anxiety (Chestnut, 1994), and social impairments such as, restrictions in their play and domestic activities, fear of underachievement, and fear of early death (Tunde-Ayinmode, 2005).

It has been recommended that the Health-related Quality of Life (HRQOL) of individuals with SCD be additionally considered as severely compromised (McClish et. al., 2005). They score significantly worse than the general population in all subscales except for mental health. They have lower HRQOL when compared to similar conditions like Cystic Fibrosis; and they fair worse than dialysis patients on bodily pain, general health, and vitality but have similarly poor assessments on physical and emotional role functions, social functioning, and mental health. Thus, improving the QOL for this population is an important health outcome (McClish et. al., 2005) and in addition to providing appropriate medical treatments, must be included as an integral part of patient management (Bhagat, Baviskar, Mudey, & Goyal, 2014).

Unfortunately, in addition to a severely compromised QOL, their overall mortality rate has been increasing by .7% per year (Lanzkron, Carroll, & Haywood, 2013). Previous research has revealed that this is likely due to lack of access to quality care, as access to quality care is associated with increased survival (Hamideh & Alvarez, 2013). This theory seems to be supported, as children with SCD who have greater access to quality care, when compared to adults with SCD, have mortality rates that have been decreasing by 3% per year (Lanzkron, Carroll, & Haywood, 2013). On the other hand, adults with SCD generally do not have sufficient access to quality care (Grosse et al., 2019) and their mortality rates are increasing by 1% per year (Lanzkron, Carroll, & Haywood, 2013). Moreover, adult mortality rates start to increase during the transitioning period where individuals with SCD move from receiving pediatric medical care to adult medical care. During this transitioning period, their mortality rates increase form 0.6 /100,000, for the 15-19-year-olds, to 1.4/100,000, for the 20-24-year-olds (Hamideh & Alvarez, 2013).

Their rising mortality rates and overall poor QOL is a devastating health inequity that exemplifies our larger societal issue related to race-based disparities with barriers to access to quality care being a primary issue. (Nelson & Hackman, 2013). For example, gaps in public and private funding for research persist (Smith, Oyeku, Homer, & Zuckerman, 2006). Cystic Fibrosis, a predominantly European American condition with a prevalence of 30,000, receives a 7.6 (2010) to 11.4-fold (2011) greater amount of funds granted towards research and drug development despite SCD, a predominantly African American condition, having the greatest genetic prevalence at 100,000 (Strouse, Lobner, Lanzkron, & MHS, 2013).

The broad use of the term “access” encompasses two parts:(1) the first relates to the quality of patient-provider relationships when the patient is receiving care (this is largely affected by providers” prior knowledge, perceptions, and communication; and (2) the second pertains to the individual”s ability to be seen, i.e., the physical presence and availability of health care services, providers, and treatment (Chen, 2007; Rouse, 2004).

Barriers to access to quality care can sometimes result from persistent racial biases and perceptions, such as the belief that African American”s don’t feel as much pain as European Americans (Hoffman, Trawalter, Axt, & Oliver, 2016) or the perception that individuals with SCD, whose pain is at minimum as bad as terminal bone cancer pain, are not suffering, are faking, or exaggerating when their use of coping and pain distraction strategies (i.e. watching TV, talking on the phone, sleeping, playing games) are witnessed (Jenerette, Pierre-Louis, Matthie, & Girardeau, 2015).

Additionally, due to the high level of pain, treatment requires large doses of opioid medications. This makes providers unfamiliar with individuals with SCD uncomfortable and hesitant to provide opioid based pain medications (Chen, 2007; Tanabe, Martinovich, Buckley, Schmelzer, & Paice, 2015). This lack of familiarity with this population coupled with racial bias leads to confounding of the differences between tolerance, dependency, and addiction. As such, some providers may believe that they are drug abusers, when they neither fit the demographics nor are more likely to be addicted than the general population. Thus, this type of barrier to access to quality care tends to lead to under-treatment of pain, delay in etiology delineation, and delay in treatment with mortal consequences (Chen, 2007).

To counteract barriers to access to quality care different models of care have been proposed. One model, the patient centered medical home, has been found to be effective at reducing the acute care utilization rates of children with SCD (Raphael et al., 2013). However, due to the severe paucity of specialists and knowledgeable providers this translates to a difficulty in establishing a medical home (de Montalembert, Guitton, & French Reference Centre for Sickle Cell Disease, 2014). A promising model that has also been proposed is the comprehensive care model. This model takes a life span and multidisciplinary approach to addressing the biopsychosocial needs of individuals with SCD, particularly those transitioning to adult medical care (Crosby, Quinn, & Kalinyak, 2015). It has been shown to reduce morbidity through early detection, preemptive actions, and by increasing patient management (Okpala et al., 2002). It is multidisciplinary as it includes nursing, nutrition, social work/case management, preventive practices, special education services, legal assistance relating to health insurance, disability benefits, and social services disciplines to provide holistic care (Baker, Crudder, Riske, Bias, & Forsberg, 2005; Grosse et al., 2009). This model is also attractive as research has proven its effectiveness in reducing mortality rates in similarly complex chronic genetic disorders, such as Hemophilia and Cystic Fibrosis (Grosse et. al, 2009).

However, this comprehensive care model has not been as effective in reducing morbidity and mortality rates of individuals with SCD as availability is lacking. Few comprehensive centers exist, and those that do exist are research based and pediatric focused. Additionally, there is no established network, nor are the few centers synchronized, making continuity of care moot (Grosse et. al, 2009). Furthermore, funding is inconsistent (Kanter & Jordan, 2015), making it even more difficult to establish and maintain such a model further reducing its availability.

Nonetheless, there are programs like Yale New Haven Health’s Adult SCD program, which through proactive interventions was able to increase access to quality care and decrease acute care utilization rates of adults with SCD in New Haven, CT. This was accomplished through bold actions by a champion physician, Dr. Roberts. Based on case studies by Rouse (2004), the actions of champion physicians can create a paradigm shift, in home institutions, where the patient and the disease matter. These physicians must be willing to change the singular problematic narrative about patients with SCD, subvert negative biases and perceptions of staff, plus encourage patient self-efficacy, ultimately leading to increased access to quality care. The Yale New Haven Health’s Adult SCD program seems to have experienced this type of paradigm shift. It was revamped through the initiative of Yale New Haven Health’s administrators who acknowledged a problem and sought a solution. Thus, they hired a transformative and progressive director, hematology-oncologists, Dr. Roberts.

The program has a dedicated adult SCD inpatient unit and outpatient clinic. It provides comprehensive medical care, which studies have found to improve the QOL, reduce medical care utilization, and length of hospital stay of individuals with SCD (Okpala et al., 2002). The program additionally takes a family centered approach to increasing patient and family self-efficacy. Care is multidisciplinary, and includes a nurse practitioner, in-patient and out-patient nurses, a psychiatrist, a hematology resident, and a chaplain. It also includes two social workers, one for inpatient and one for outpatient care, who form a bridge between the health care system and psychosocial issues. Through Dr. Robert’s great knowledge, understanding, and compassion for this patient population, he and his staff have been able to provide pain control, increase access to quality care, improve the QOL of his patients, and increase provider availability (Rousseau, n.d.).

In 2005, Shankar et al. identified that when compared to Tennessee’s general black population, individuals with SCD enrolled in the state’s Medicaid program had higher: race and age specific mortality rates, ED utilization rates (2-6 times higher), and hospitalization rates (7-30 times higher). This study revealed a pattern of high medical care utilization among the SCD population and an urgent need to produce interventions to decrease mortality rates. However, it was restricted to Medicaid patients and to the state of Tennessee which limited generalizability.

Brousseau et al. (2010), taking into consideration limitations to generalizability conducted a population-based study, across eight geographically diverse states (Arizona, California, Florida, Massachusetts, Missouri, New York, South Carolina, and Tennessee). They investigated acute care utilization rates, as well as 30-day and 14-day re-hospitalization rates which they used as indicators for the quality of care provided. Their results substantiated previous studies in that individuals with SCD had higher acute care utilization rates. They also found that individuals with SCD 18-30-years old, who were transitioning from pediatric to adult medical care, had the highest rates of acute care encounters and rehospitalizations. Such high acute care utilization rates are of concern as Hunt & Sharman (2010), found that high rates of acute care utilization and early readmissions are correlated with increased mortality rates.

Wolfson et al. (2012), investigated sociodemographic predictors of ED utilization rates in California and found that age, insurance type, and location were factors related to increased acute care utilization rates. Specifically, they found, similar to Brousseau et al. (2010), that adults, ≥ 21 years of age, had higher acute care utilization rates. They also found that those with public insurance and those who lived a greater distance away from a comprehensive medical care also had higher acute care utilization rates. Their findings suggested that these factors may act as barriers to access to quality care.

However, to better identify barriers to access to quality care and health care utilization rates, population-based surveillance data is the recommended source. Nevertheless, such a surveillance system is lacking for SCD (Hoots, 2010; Grosse, James, Lloyd-Puryear & Atrash, 2011). In recognition of this deficit, from 2010-2012, the Center for Disease Control and Prevention (CDC) developed the Registry and Surveillance in Hemoglobinopathies (RuSH) in partnership with seven states (Hoots, 2010). However, as the resultant data was limited to those participating states, previous research on health care utilization continued to rely on administrative data, such as hospital discharge records and Medicaid records. These secondary sources of data are limiting as they are unlinked, may be biased and may miss healthier individuals. Data from these sources leads to an estimation of population prevalence and not a true count (Paulukonis, 2014). Thus, making it difficult for public health initiatives to identify individuals with SCD and to monitor their health care utilization rates and health outcomes (Hulihan, 2015).

To strengthen confidence in the utilization of administrative data for the assessment of health care utilization rates of individuals with SCD, different methods have been employed. The needs assessment conducted by Schlenz, Boan, Lackland, Adams & Kanter (2016), in South Carolina evaluated statewide administrative data. Similar, to Wolfson et al. (2012), they reached the conclusion that age, insurance type, and location were predictive factors of acute care utilization rates. More importantly, they highlighted how administrative data could provide the necessary information to assess health care utilization rates for individuals with SCD. Furthermore, the study conducted by Benenson, Jadotte & Echevarria (2017), involved the use of systematic reviews to identify factors that influence the health care utilization rates of adult SCD. While the study by Brennan-Cook et.al (2018), identified case managers, who play key roles in addressing the needs of individuals with SCD, as a potential access to quality care study topic.

Individuals with sickle cell disease are a forgotten group of people who live physical and mentally debilitated lives. As a society we cannot continue to ignore their increasing death rates. Nor can we continue to relegate them to those who suffer in silence. This research can help to fill the gap in research, provide data, and provide an understanding about factors that positively influence health outcomes. It will provide information about systematic and multilevel factors that can be utilized to promote public health, promote preventive strategies in their treatment and management, and in the development of interventions.

Public health professionals, physicians, nurses, local, state, and national politicians/policy developers and advocates, community organizations, patients, family, and friends will have key information that can be utilized in the short and long-term towards positive change. It will aid in increasing funds towards research ultimately helping to close the gap in this health inequity. It will also provide the first step to the development of a cost-effective framework for other communities to use in their community needs assessments, identify policies that strengthen the community eventually creating a social environment that allows the community to meet the needs and provide the necessary resources to their members with SCD.

Therefore, the goal for this project was to conduct a mixed method needs assessment with an objective to conduct an initial analysis of administrative data to assess barriers and opportunities as it relates to access to quality care. The other objective was to supplement the quantitative analysis with qualitative data, in order to gain the perspectives of key informants. The qualitative data would present deeper and more contextualized understanding of the unique needs of individuals with SCD and provide solutions to address barriers that they face. The needs assessment is an initial step towards the development of a community engaged SCD comprehensive center.

By the end of this project the researcher hopes to satisfy the general elements of public health, fulfill elements of the public health framework for rare blood disorders (Grosse, James, Lloyd-Puryear, & Atrash, 2011), and fulfill the CDCs Healthy People 2020 goals by producing surveillance data, providing a local sampling frame that can be used for future research, and produce data that can be used to inform interventions to increase access to quality care. The researcher ultimately hopes to start a conversation and increase awareness in the community, in medicine, and public health about the most prevalent genetic disorder and a health disparity that was the foundation of molecular science and is a harbinger of societal neglect and inequity.

## Method

### Hypothesis

**Ho1.** There is no difference in emergency department usage rates, in-patient hospital admission rates, length of stay rates, and outpatient visit rates between age groups.
**Ha1.** There is a difference in emergency department usage rates, in-patient hospital admission rates, length of stay rates, and outpatient visit rates between age groups.
**Ho2.** There is no difference in emergency department usage rates, in-patient hospital admission rates, length of stay rates, and outpatient visit rates between payer types.
**Ha2.** There is a difference in emergency department usage rates, in-patient hospital admission rates, length of stay rates, and outpatient visit rates between payer types.

### Data source

De-identified administrative data for emergency department encounters, hospitalizations (inpatient encounters), in patient lengths of stay (LOS), outpatient visit encounters, payer type, age, and gender were obtained from the University of Florida (UF) Health’s Integrated Data Repository (IDR). Data with primary or secondary discharge diagnosis of Sickle Cell Disorders D57s, based on the International Classification of Disease Tenth Revision (ICD 10-CM), from September 01, 2002, to October 01, 2015; and 282.60s, based on ICD 9-CM, from October 01, 2015, to June 01, 2019, were included (See Appendix A). Patients who were not assigned an age at encounter were excluded.

Qualitative data of perspectives were obtained from semi-structured interviews of 10 key informants who have provided care and advocated on behalf of this population at the institutional, community, and policy levels. Snowball sampling was used to recruit key informants in Alachua County, FL, starting with a community partnership with the local Sickle cell Disease Associations of America (SCDAA). Leaders in the organization were interviewed and they recommended providers that were subsequently interviewed. A purposeful sampling was used to recruit key informants at Yale New Haven Health’s Adult Sickle Cell program, where providers and staff were interviewed at the hospital and clinic.

Interviews lasted from 45 minutes to one hour and a half and all interviewees received questions related to their perspective on: barriers to access to quality care, on the QOL of individuals with SCD, psychosocial barriers and needs, and their recommendations for increasing access to quality care. Providers received additional questions related to patient management, while community interviewees received additional questions related to social barriers and what individuals with SCD have indicated to them as needs. Key informants at Yale were additionally asked about the successes of their program, challenges experienced, opportunities, and lessons learned from implementing their program. Key informants were interviewed and recorded. Eight interviews were recorded with responses written and two interviews were not recorded with responses only written.

### Variables

#### Age

Age of individual was determined as age at each encounter. Age was further stratified in the following groups: 0-9, 10-17, 18-30, 31-45, 46-64, and 65+, consistent with previous SCD care utilization research.

#### Payer

Payer (used as a proxy for socioeconomics) was determined as payer listed at each encounter. This included Medicaid, Medicaid HMO, Medicare, CMS, and Federal categorized as Public; Blue Cross, Commercial, and Managed Care categorized as Private; Self-pay categorized as Self-Pay; and Jax Charity, Workers’ comp, and other categorized as Other.

### Data analysis

Descriptive analyses were completed in IBM® SPSS® Statistics version 25. Frequency, proportions, contingency-tables, Pearson Chi square, and P-values for medical care encounters per patient were stratified by age and payer type, for ED visits, hospitalizations, outpatient visits and total encounters. Mean and standard deviation for LOS for medical care encounters per patient were stratified by age and payer.

A thematic framework analysis was used to analyze the qualitative data. Recordings were re-listened to and written responses were reread for emerging patterns. Themes that emerged in response to the semi-structured interview were recorded in a themes table. Words and phrases were coded as a theme if they appeared multiple times during an interview, if it occurred in a separate interview, or if it provided a deeper meaning to perspectives of key informants as it relates to the needs of individuals with SCD.

## Results

The final data set consisted of n=27,932 total care encounters that were stratified by age and payer type. The majority were outpatient encounters at n=18,582 (66.5%), while n=5,748 (20.6%) were inpatient, and n=3,601 (12.9%) were ED encounters. Most encounters consisted of the 18-30-year-olds (n=8055), followed by the 31-45-year-olds (n= 5779), and then the 46-64-year-olds (n=4405). The 18-30-year-olds had the greatest proportion of ED encounters at 36%, followed by the 31-45-year-olds at 21%, then the 46-64-year-olds at 19%. The 18-30-year-olds had the greatest proportion of hospitalizations at 32%, followed by the 31-45-year-olds at 22 %, then the 46-64-year-olds at 17%. The 18-30-year-olds also had the greatest proportion of outpatient visit encounters at 28%, and again followed by the 31-45-year-olds at 22 %, and the 46-64-year-olds at 16% (See Figure 1).

**Figure 1.**
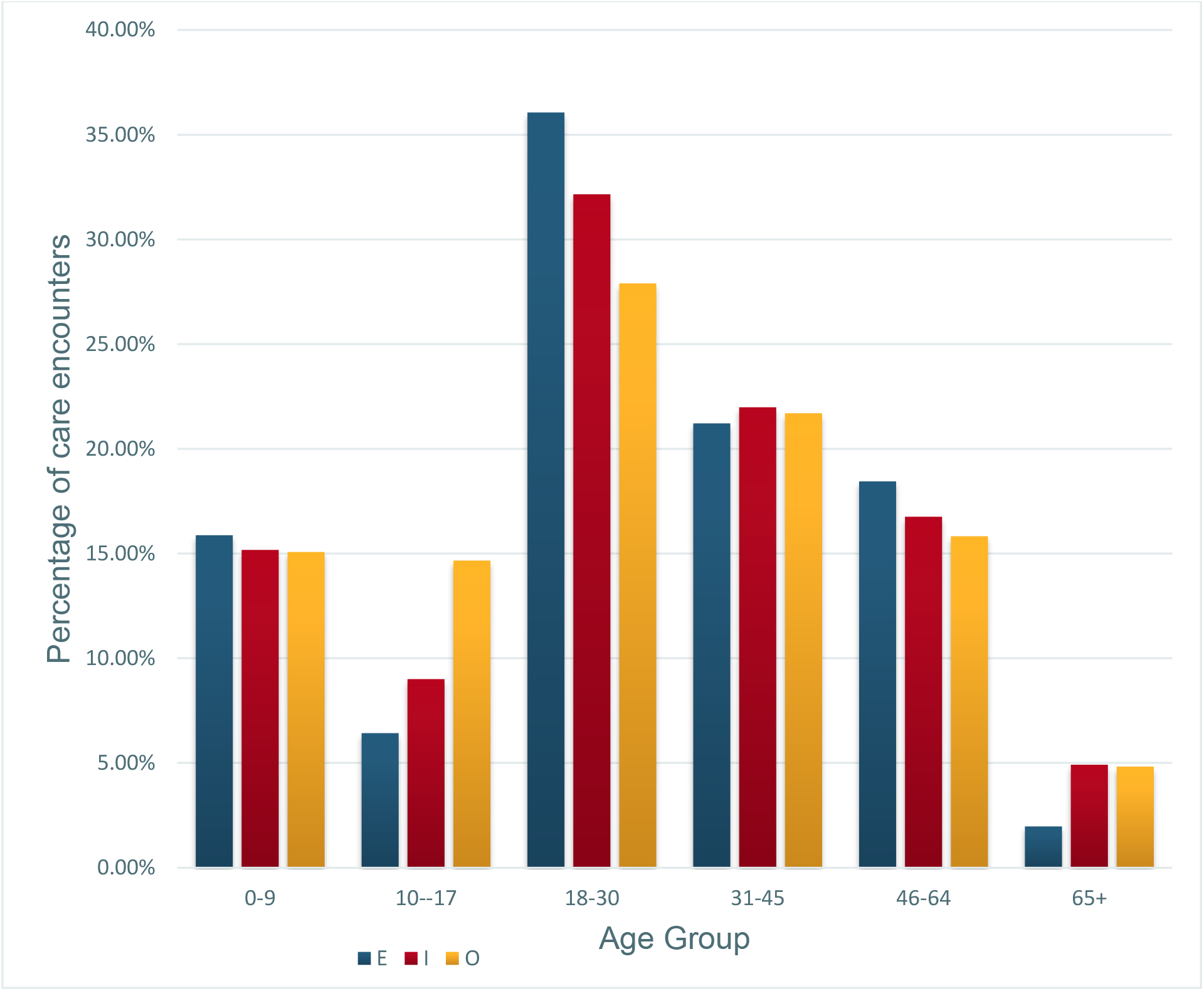
UF Health Care utilization for patients with Sickle Cell Disease from 2012-2019.E=emergency department utilization, I=hospitalization, O=outpatient clinic visit.

For inpatient LOS, the overall average LOS by payer was .74 ± 3.93 days. The overall average LOS by age was similar at .71 ± 3.84 days. LOS stratified by age shows that the 65+-year-olds had the longest average LOS at 1.04 ± 3.5 days, followed by the 45-64-year-olds at .83 ± 4.0 days, and then the 31-44-year-olds at .80 ± 3.8 days. The 0-9-year-olds had a LOS of .78 ± 6.5 days, while the 18-30-year-olds had a LOS of .71± 2.6 days. The 10-17-year-olds had the shortest average LOS at .47 ± 2.5 days (See Figure 2).

**Figure 2.**
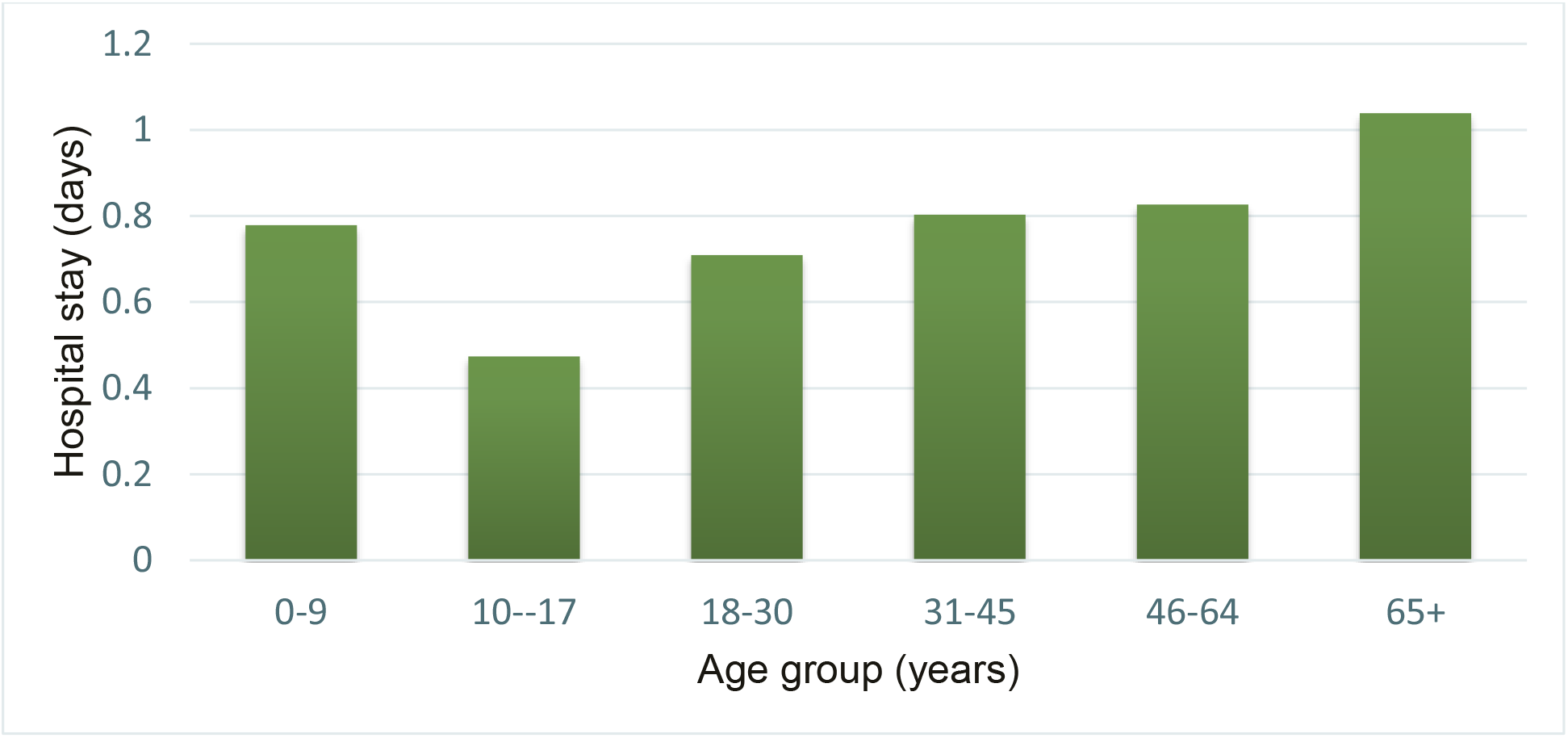
Age group Average Length of Hospital stay (LOS).

Most had Public insurance (79.92%), followed by Private (15.51%), then Self-pay (2.71%), with the least being Other (1.85%) (See Figure 3). Those with public insurance had the greatest proportion of ED encounters at 74%, followed by those with private insurance at 17%, with self-pay at 6%, and other type of insurance at 3%. Persons with public insurance also had the greatest proportion of hospitalizations at 81%, followed by those with private insurance at 18%, with similar proportions for other type of insurance and self-pay at 2.47% and 2.44% respectively. Individuals with public insurance also had the greatest proportion of outpatient visit encounters at 82%, followed by those with private insurance at 15%, with similar proportions for self-pay and other type of insurance at 1.63% and 1.52% respectively (See Figure 4). Those with other type of insurance had the longest average LOS at 1.1 ±3.6 days, followed by those with public insurance at .73 ± 3.9 days, then private insurance at .6 ± 3.7 days, with self-pay with the lowest average LOS at .57 ± 2.3 days.

**Figure 3.**
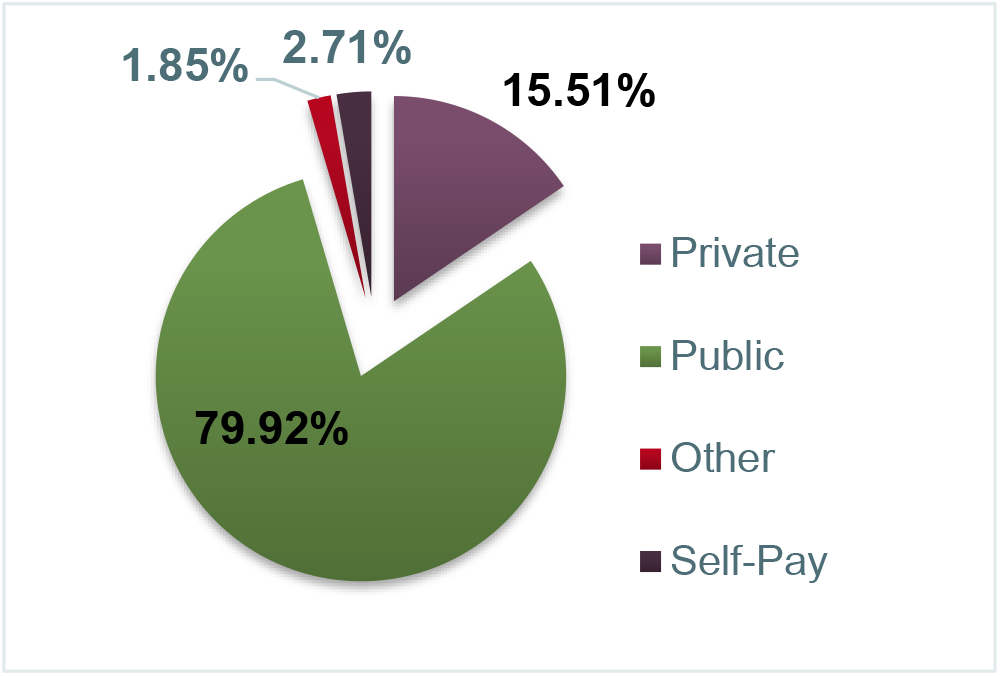
Payer Mix

**Figure 4.**
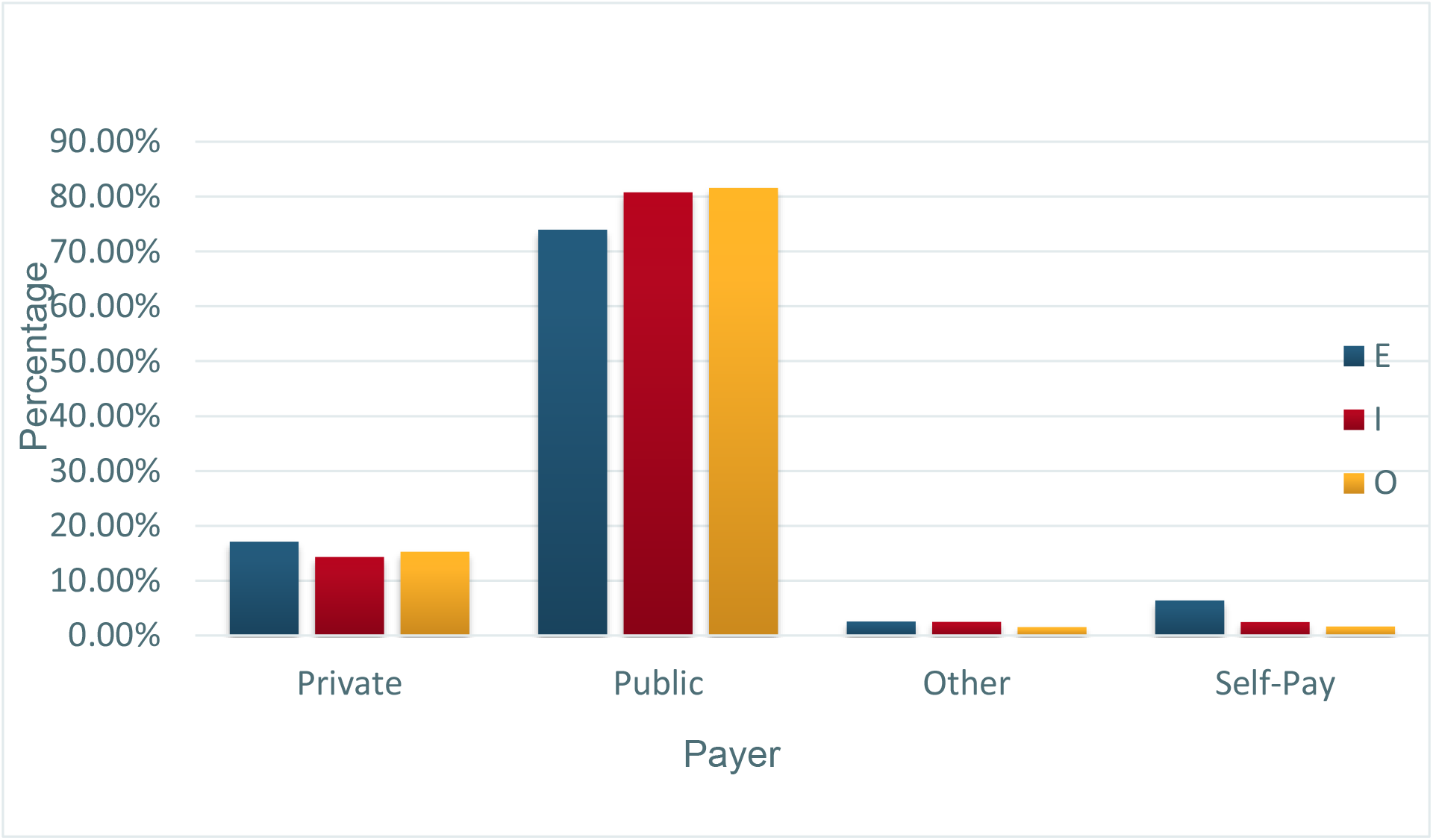
UF Health Care Utilization by Payer type for patients with Sickle Cell Disease from 2012-2019. E=emergency department utilization, I=hospitalization, O=outpatient clinic visit

For encounters stratified by age and payer the p-value was <.001. The degree of freedom was 12, the minimum expected count was 146.33, and none of the cells had an expected count of less than 5. Based on these results we can conclude that there is enough evidence to reject the null hypothesis and accept the alternative hypothesis that there is a statically highly significant association between age and payer type with health care utilization encounters.

Themes that emerged from the key informant interviews included: champion, transition of care, pain management, bias, patient and family education, provider knowledge, social worker, multidisciplinary/comprehensive care, mental health, education, employment.

## Discussion

The aim of the mixed method study was to primarily assess the health care utilization rates of individuals with SCD in Alachua County, FL and to investigate possible barriers and opportunities to access to quality care. The current study additionally aimed to identify SCD specific social determinants of health and identify constructs that may be used for future research. With the ultimate goal that the results may be used to develop interventions to improve the QOL and reduce the mortality rates of individuals with SCD.

A quantitative analysis of administrative data was used to evaluate if there was a difference in health care utilization rates and LOS when stratified by age and payer type. Health care utilization rates were selected as research variables as previous research has shown that it is associated with increased mortality rates (Lanzkron, Carroll, & Haywood, 2013). As state population-based surveillance was lacking, administrative data was used instead. Administrative data was selected as previous studies have proven their utility in assessing the acute care utilization rates of individuals with SCD at the state and institutional levels (Shankar et al, 2005; Schlenz, Boan, Lackland, Adams & Kanter, 2016). Based on the literature search, a qualitative approach was taken as it is often not combined with quantitative studies when assessing access to quality care and health care utilization rates of individuals with SCD. This current study, through semi-structured interviews of key informants, fills in that gap. A qualitative method was, additionally, combined to gain a deeper understanding of barriers to care, the psychosocial needs of individuals with SCD, and to identify potential solutions from individuals with experience providing care and advocating for this patient population.

The findings of the study are consistent with those of Brousseau et al. (2010), as the distribution of the encounters indicates a relationship between health care utilization with age and payer type. The results suggest that the 18-30-year-olds utilize health care services the most, while the 10-17-year-olds utilize health care services the least, when we excluded those who are 65+ years of age. However, the LOS was lowest for the 18-30-year-olds and the 10-17-year-olds, indicating that there may be some type of barrier to gaining access to quality care, particularly preventive care.

Coinciding with the quantitative analysis, the semi-structured interviews also identified the 18-30-year-olds as high health care utilizers, suggesting that they may be experiencing greater barriers to accessing care. Moreover, the semi-structured interviewees indicated that this age group would benefit significantly from a transition of care program that starts earlier and extends further into adulthood. Their responses correlate with Bemrich-Stolz, Halanych, Howard, Hilliard & Lebensburger‘s (2015), study recommendation for the need to implement a formalized transitioning program to help mitigate barriers to access to quality care for the 18-30-year olds with SCD, as they transition through care.

Barriers to access to quality care leads to significant physical and psychosocial burdens that severely compromises their HRQOL, overall QOL, and wellbeing. Based on the results of this study and when compared to contemporary literature related to acute care utilization rates, we can infer that there is a barrier to access to quality care and an urgent need to increase access to quality care for adults with SCD in Alachua County, FL. Individuals with SCD who are transitioning from pediatric to adult medical care, adults with SCD, and those with public insurance are viable targets for interventions and further study.

Confidence in the results of this study are strengthened as the study utilized an objective and comprehensive database like IDR, which is routinely monitored and in compliance with IRB protocols, for administrative data. The use of administrative data minimizes researcher bias and strengthens the external validity of the results (Harron et al., 2017). Furthermore, to better capture patients and patient encounters a very broad and inclusive coding systems of diagnosis were used. Specifically, the study utilized both the DSM9 and DSM10 codes to identify the patients. The study is furthered strengthened by its use of administrative data as previous research has concluded that this type of data source is a viable source for assessing health care utilization rates of individuals with SCD in the absence of population-based surveillance data (Schlenz, Boan, Lackland, Adams & Kanter, 2016). The sample size of encounters was large which increases statistical power. The study collects data across an extensive time period which aids to effectively track and trend patterns of healthcare utilization rates.

On the other hand, use of administrative data is also limiting as data may not be linked accurately. The data may be misreported, misclassified, or missing. Thereby, reducing the reliability of the data (Harron et al., 2017). Use of a secondary data source is also a limitation as the patients’ medical records were not individually investigated by the researcher to analyze if the same encounters were counted multiple times. Some encounters were missing an age and had to be excluded. This exclusion might have changed the results of the study had they been included. However, as the number excluded was small, in comparison to the sample size, exclusion of those encounters are not likely to significantly change the results.

The results are not generalizable to the larger SCD population as this was performed on a limited data sample of patients in the UF Health system. UF is a prominent medical institution with a positive history of providing care so results may be skewed. Results may also vary depending on location. Other settings such as non-academic or community-based health centers or clinics without an attached outpatient clinic may have higher ED utilization rates. Results may also vary for more urban or rural settings. Though Gainesville, FL is not considered to be a rural city it is surrounded by rural communities which may skew the results towards higher utilization.

## Implications

This research provides initial surveillance data and a sampling framework that can be used for further research. The results can inform the design of interventions and contribute to larger projects like One Florida’s project for SCD. It also provides the first step towards the development of a community engaged SCD program, as it generated data from a needs assessment and resulted in collaborations with a community organization. SCD is a disparity that has been declared a public health priority by the CDC (Center for Disease Control and Prevention (n.d.). Yet, though it is a public health condition, it is not often recognized as such (Grosse, James, Lloyd-Puryear & Atrash, 2011; Yusuf et al., 2011). Unfortunately, the combination of multiple and complex factors of pain, implicit bias, provider and patient knowledge, stigma, and high acute care utilization, cause individuals with SCD to experience persistent barriers to access to quality care, higher mortality rates, and poorer QOL (Brennan-Cook, Bonnabeau, Aponte, Augustin & Tanabe, 2018).

Thus, this project attempts to raise awareness across all professions from public health to medicine. Hoping that if questions are raised and conversations are started it will encourage academics to consider SCD in their research endeavors. SCD is the most prevalent genetic condition in this nation and globally, yet it has some of the lowest amount of funding and research when compared to similar chronic genetic disorders and rare blood disorders (Lobner, Lanzkron, & Haywood, 2013; Grosse, James, Lloyd-Puryear & Atrash, 2011). If we, in academia, can draw attention and make a call then funding and capacity will likely increase. As a result, researchers and interventionists may be able to freely engage in innovative research to identify factors; and to design and implement effective evidence-based interventions.

## Data Availability

Secondary data sets available through University of Florida's Integrated Data Repository. Anonymized primary data set available from researcher.

https://idr.ufhealth.org/

## Appendix A SICKLE CELL DISEASE DIAGNOSIS

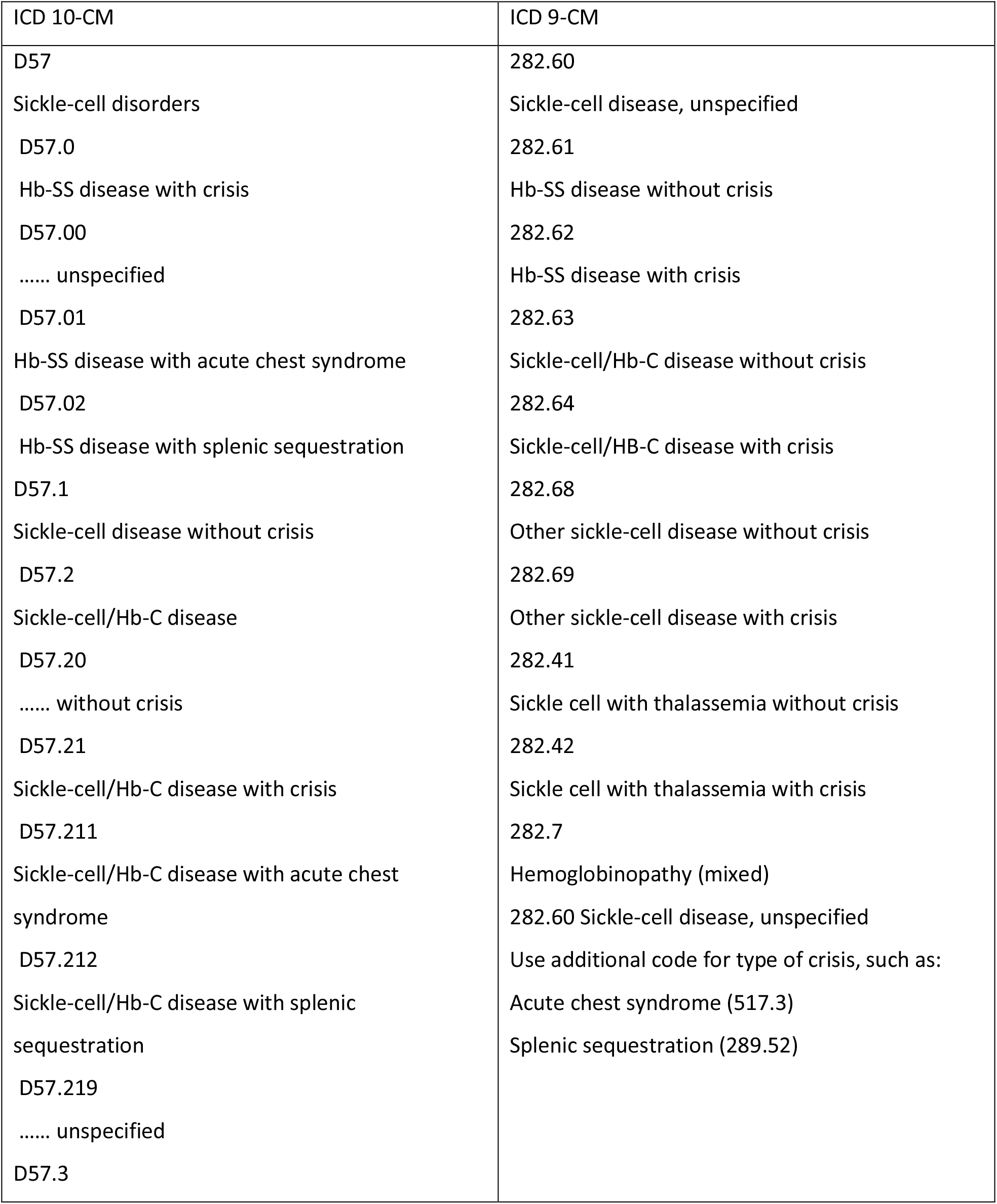
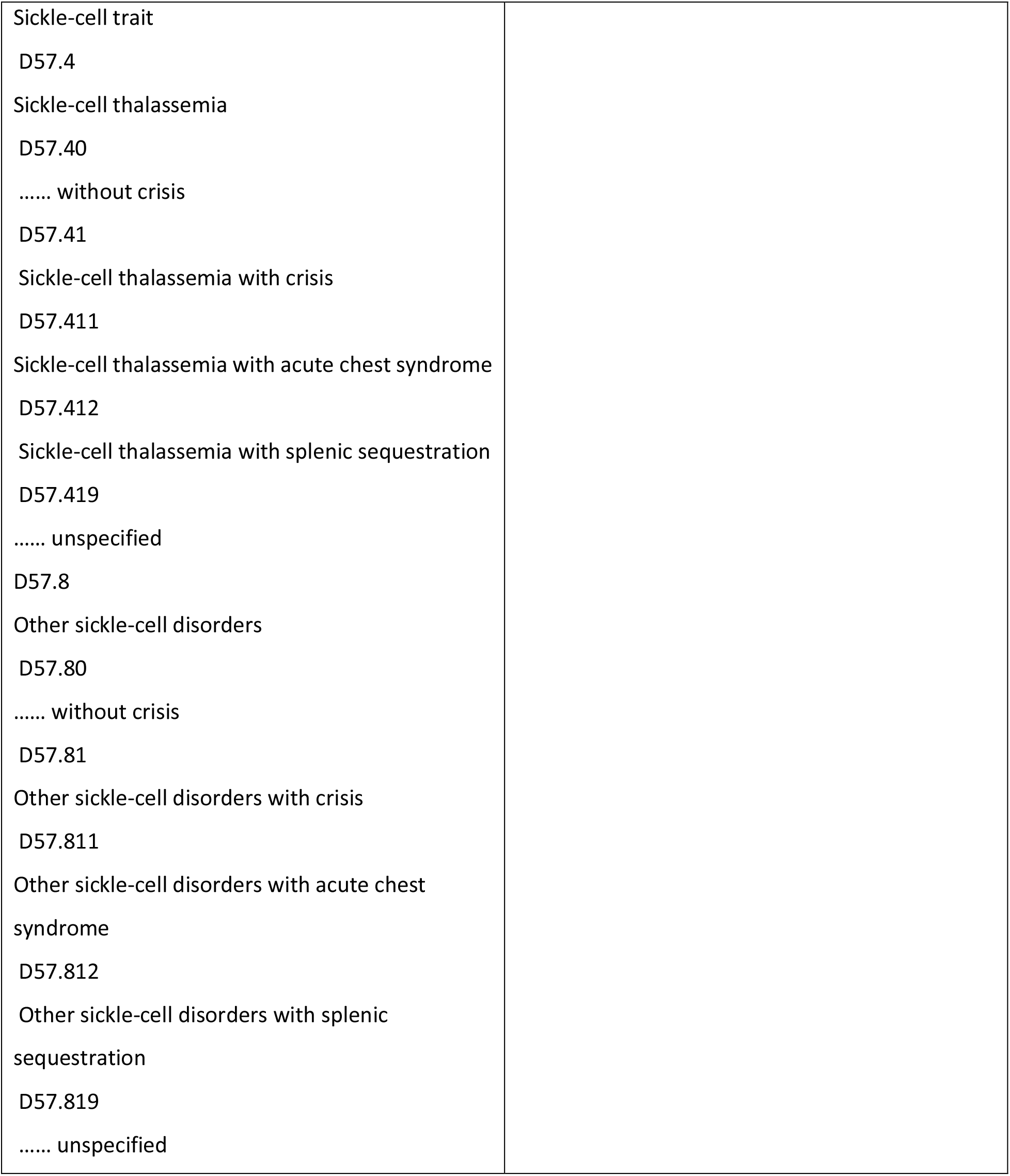

SICKLE CELL DISEASE DIAGNOSIS
| ICD 10-CM | ICD 9-CM |
| --- | --- |
| D57 | 282.60 |
| Sickle-cell disorders | Sickle-cell disease, unspecified |
| D57.0 | 282.61 |
| Hb-SS disease with crisis | Hb-SS disease without crisis |
| D57.00 | 282.62 |
| ..... unspecified | Hb-SS disease with crisis |
| D57.01 | 282.63 |
| Hb-SS disease with acute chest syndrome | Sickle-cell/Hb-C disease without crisis |
| D57.02 | 282.64 |
| Hb-SS disease with splenic sequestration | Sickle-cell/Hb-C disease with crisis |
| D57.1 | 282.68 |
| Sickle-cell disease without crisis | Other sickle-cell disease without crisis |
| D57.2 | 282.69 |
| Sickle-cell/Hb-C disease | Other sickle-cell disease with crisis |
| D57.20 | 282.41 |
| ..... without crisis | Sickle cell with thalassemia without crisis |
| D57.21 | 282.42 |
| Sickle-cell/Hb-C disease with crisis | Sickle cell with thalassemia with crisis |
| D57.211 | 282.7 |
| Sickle-cell/Hb-C disease with acute chest syndrome | Hemoglobinopathy (mixed) |
| D57.212 | 282.60 Sickle-cell disease, unspecified |
| Sickle-cell/Hb-C disease with splenic sequestration | Use additional code for type of crisis, such as: |
| D57.219 | Acute chest syndrome (517.3) |
| ..... unspecified | Splenic sequestration (289.52) |
| D57.3 |  |
| Sickle-cell trait<br>D57.4<br>Sickle-cell thalassemia<br>D57.40<br>..... without crisis<br>D57.41<br>Sickle-cell thalassemia with crisis<br>D57.411<br>Sickle-cell thalassemia with acute chest syndrome<br>D57.412<br>Sickle-cell thalassemia with splenic sequestration<br>D57.419<br>..... unspecified<br>D57.8<br>Other sickle-cell disorders<br>D57.80<br>..... without crisis<br>D57.81<br>Other sickle-cell disorders with crisis<br>D57.811<br>Other sickle-cell disorders with acute chest syndrome<br>D57.812<br>Other sickle-cell disorders with splenic sequestration<br>D57.819<br>..... unspecified |  |

## Notes

### Competing Interest Statement

The authors have declared no competing interest.

### Funding Statement

No external funding received.

### Author Declarations

University of Florida Institutional Review Board Gainesville Health Science Center/Jacksonville Health Science Center (IRB-01) Committee. Approved.

